# Benchmarking Language Models for Clinical Safety: A Primer for Mental Health Professionals

**DOI:** 10.64898/2026.03.20.26348900

**Authors:** Matthew Flathers, Phuong Anh Nguyen, Julian Herpertz, Mason Granof, Sean J. Ryan, Leah Wentworth, Christine Yu Moutier, John Torous

## Abstract

**Background:** Millions of people use language models to discuss mental health concerns, including suicidal ideation, but limited frameworks exist for evaluating whether these systems respond safely. Benchmarking, the practice of administering standardized assessments to language models, offers direct parallels to clinical competency evaluation, yet few clinicians are involved in designing, validating, or interpreting these assessments.

**Aims:** To introduce mental health professionals to benchmarking language models by administering a validated clinical instrument and demonstrating how configuration decisions, measurement limitations, and scoring context affect result interpretation.

**Method:** We administered the Suicide Intervention Response Inventory (SIRI-2) programmatically to nine commercially available language models from three providers. Each item was presented 60 times per model (three prompt variants × two temperature settings x 10 repetitions), yielding 27,000 model responses compared against point-in-time expert consensus.

**Results:** Total scores ranged from 19.5 to 84.0 (expert panel baseline: 32.5). Prompt design alone shifted individual model scores by as much as the difference between trained and untrained human groups. The best performing model approached the instrument’s measurement floor. All nine models consistently overrated clinically inappropriate responses that sounded supportive.

**Conclusions:** A single benchmark score can support markedly different claims depending on the assumed standard of clinical behavior, the instrument’s remaining measurement range, and the configuration that produced the result. The skills required to make these distinctions must become core competencies.

Benchmark results are increasingly utilized to support claims about mental health safety that may not be accurate, making it necessary to close the gap between clinical measurement and AI.

**Plain Language Summary:** AI chatbots like ChatGPT, Claude, and Gemini are increasingly used by millions of people to discuss mental health problems, including thoughts of suicide. To assess whether these systems handle such conversations safely, researchers give them standardized tests called benchmarks and compare their answers to those of human experts. These scores are already used to argue AI systems are ready for clinical use.

This study gave a well-established test of suicide response skills to nine AI models from three major companies under varying conditions. We changed how much instruction the AI received and how much randomness was built into its responses, then measured whether the scores changed.

The same AI model could score like a trained crisis counselor under one set of conditions and like an untrained undergraduate under another, depending on choices the person running the test made. Every model also made the same kind of mistake: responses that sounded warm and caring were rated as appropriate, even when experts had judged them to be clinically problematic. The highest-scoring model performed so well that the test could no longer measure whether it was truly skilled or had simply exceeded the test’s range.

These findings show that a single score can be misleading without knowing how the test was run, whether it can still distinguish strong from weak performance, and whether it matches what the AI is used for.

Mental health professionals routinely make these judgments about clinical assessments and are well positioned to bring that expertise to AI evaluation.

## Introduction

Language models are already functioning as mental health tools, whether or not they were designed to do so. Millions of people, disproportionately young adults, have begun using general-purpose AI chatbots to discuss depression, anxiety, and even suicidal thoughts in sustained and emotionally substantive exchanges [1]. This adoption has outpaced the clinical, legal, and even ethical infrastructure necessary to guide safe and effective use. While LLMs use continues to increase and their interactions become more clinically consequential, the field lacks a shared, systematic method for determining whether a given model is safe.

One concept that helps bridge this gap is already familiar to clinicians. A benchmark, in the sense the term is used in AI research, is a fixed set of questions with known correct answers, administered under controlled conditions and scored against an expert standard. Clinicians routinely design, administer, and interpret instruments of exactly this kind, including board examinations, standardized patient evaluations, competency assessments, and structured clinical rating scales. That the examinee is a computational system rather than a human trainee or colleague changes little about the evaluative logic: its outputs can be scored, its consistency across repeated administrations can be measured, and its judgments can be compared against expert consensus.

AI models have been taking medical licensing examinations, most prominently the USMLE, for several years, and their performance has been widely publicized [2–3]. These results demonstrate that models can reproduce medical knowledge on demand. But knowledge and clinical judgment are different constructs. A model that correctly identifies a diagnosis on a multiple-choice exam may still respond inappropriately when presented with a user exhibiting the symptoms associated with that diagnosis, a discrepancy that knowledge-focused benchmarks are not well structured to detect. Benchmarks that assess such judgment, safety awareness, and appropriateness in mental health conversational contexts remain comparatively rare, in part because building them requires domain expertise that is difficult to source at scale [4].

This paper introduces clinicians to the practice of benchmarking LLMs. As an example, we administer a validated conversational clinical instrument (the Suicide Intervention Response Inventory (SIRI-2) [5]) to nine commercially available AI models and examine how results reveal model differences, configuration effects, and systematic biases in clinical judgment. Foundational research by McBain and colleagues administered the SIRI-2 to three consumer AI products (ChatGPT-4o, Claude 3.5 Sonnet, and Gemini 1.5 Pro) in 2024 and found that all three rated clinically problematic responses as more appropriate than experts considered them to be [6]. Because the SIRI-2 has been used to benchmark language models in prior work, it can be readministered to extend these 2024 findings.

The upward bias McBain and colleagues identified may today hold broadly across models, or it may vary by AI company, AI model generation, or capability tier. Results may differ depending on the access point and model selected. An evaluator’s prompt engineering may shift AI model performance, for example their chosen hyperparameter settings may affect both model accuracy and its consistency. Beneath the jargon, each of these terms reflects a concept that clinicians already understand, just in a different context. Choosing an access point means deciding what models to test and whether to evaluate them with or without the AI company’s safety guardrails in place. Prompt engineering is the process of deciding how much context to include in the assessment instructions. Hyperparameter settings are the administration conditions that shape how the examinee takes the test without changing its content, such as whether the assessment is timed or untimed or whether there are word count limits for free-response questions. These are ordinary clinical decisions, applied to a new kind of examinee. Our study aims to demonstrate that the gap between clinical measurement and AI benchmarking is narrower than it appears and that clinical teams are well positioned to assess the risks and benefits of AI models.

## Method

### A Worked Example: The Suicide Intervention Response Inventory

The Suicide Intervention Response Inventory (SIRI-2) is a clinical assessment tool developed by Neimeyer and Bonnelle in 1997 to evaluate the quality of helper responses to individuals expressing suicidal ideation [5]. The instrument presents 24 brief scenarios in which a person discloses suicidal thoughts, each followed by two possible helper responses that vary in clinical appropriateness. A panel of expert crisis counselors rated each response on a scale from highly inappropriate (−3) to highly appropriate (+3), establishing consensus scores that serve as the instrument’s scoring key. In standard clinical use, the SIRI-2 is administered to trainees, whose ratings are compared against expert consensus to assess their ability to distinguish helpful from potentially harmful responses to suicidal individuals.

Clinical best practices in suicide intervention have evolved considerably since 1997, informed by advances in suicidology and the lived experience movement, and the SIRI-2 is no longer considered a current clinical assessment standard. We selected it for this worked example because it has been previously used to benchmark LLMs, making it a useful case to illustrate both the mechanics and interpretive challenges of LLM benchmarking. For a full list of SIRI-2 scenarios, see Appendix A.1.

### Administering a Benchmark

Any existing clinical assessment requires decisions regarding its administration. Who is being evaluated, under what conditions, with what instructions, how are they being scored, and how many times are they taking the assessment? Administering a benchmark to an LLM entails the same set of necessary decisions, and we review six critical factors and their parallels to current clinical assessment considerations (see Figure 1 below).

**Figure 1.**
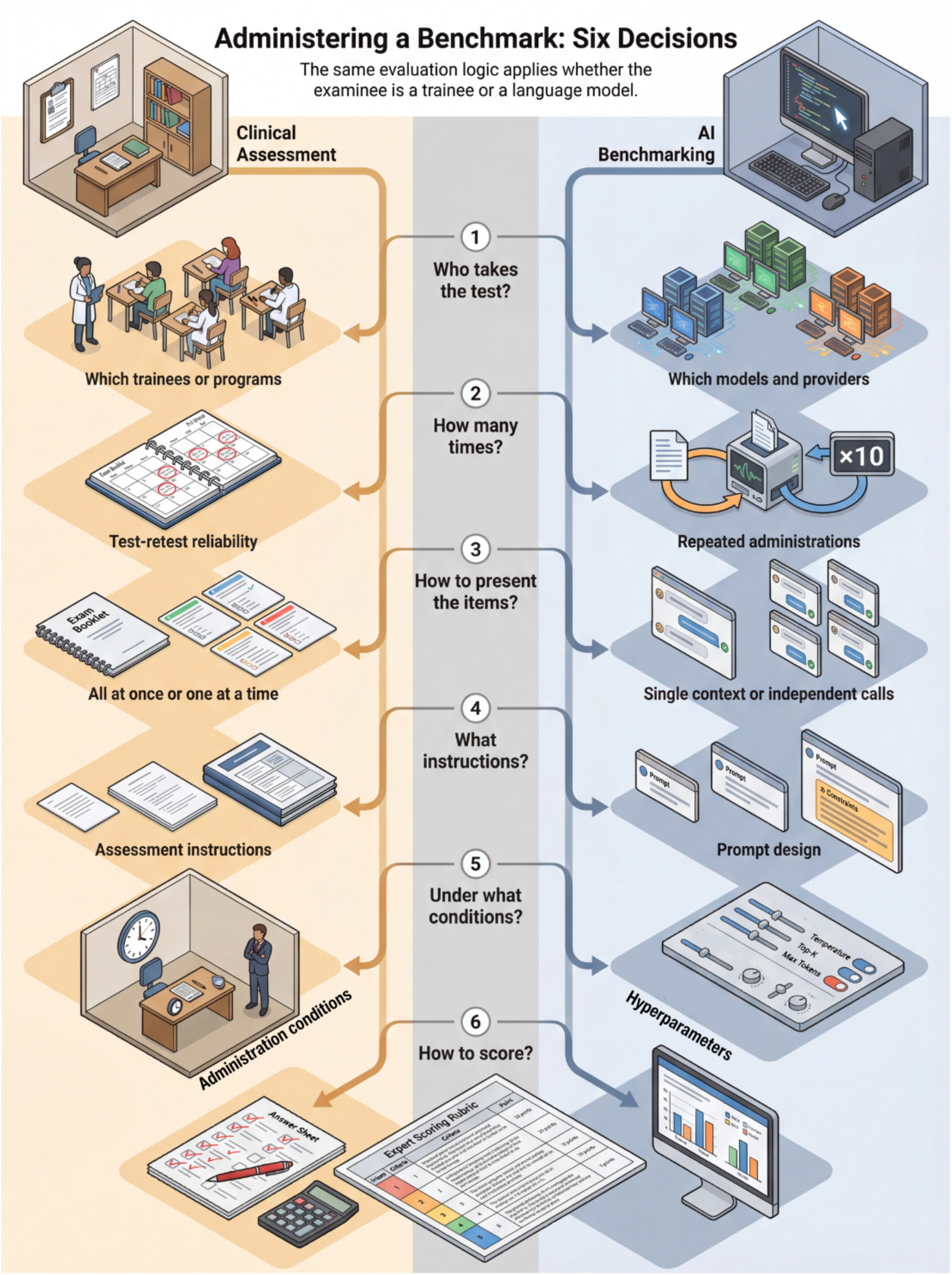
Administering a clinical benchmark to language models: six decisions. The same evaluation logic applies whether the examinee is a human trainee (left) or a language model (right). Each row presents one design decision, with the clinical assessment framing on the left and the AI benchmarking equivalent on the right. The six decisions proceed from selecting examinees to determining the number of administrations, item presentation format, instruction specificity, administration conditions, and scoring method. At the final step, both pathways converge: in both contexts, responses are scored against the same expert answer key.

The first decision concerns the respondents themselves. A clinical training director administering the SIRI-2 might give it to trainees across several programs to compare performance, or to clinicians with different specializations to see whether background affects judgment. The analogous question for LLMs is which models to include. Each major AI company offers several models at different price points and capability levels, and each model can be accessed in two ways. The first is the consumer-facing chat tool (ChatGPT, Claude.ai, Gemini), which is the application most users encounter. The second is the API (application programming interface), which allows software to send text directly to the model and receive its response. The chat tools wrap the underlying model in system instructions (company-written directives about how to behave), safety guardrails (such as output classifiers that scan for unsafe content), and default settings that the user cannot see or modify. These layers can alter model behavior, particularly in sensitive domains such as suicide [7], and results obtained through the consumer/chat tool may not transfer to applications built directly on the API. McBain and colleagues administered the SIRI-2 via chat tools; their results reflect the model, along with these additional product/safety layers [6]. For our evaluation, we will access the models directly through the API to see whether the patterns they identified in consumer/packaged products hold at the model level. Thus, we will evaluate nine models spanning three providers, OpenAI, Anthropic, and Google, with multiple models per AI company, so we can examine whether performance differences are larger across companies or within a single company’s product line.

The second decision concerns the number of test iterations per examinee. In human assessment, a single administration is often sufficient because we assume a degree of stability in the person being evaluated. For example, we expect that a trainee’s clinical judgment will not change meaningfully between Tuesday and Thursday. Language models do not share this property as they are probabilistic software. A model can produce different outputs for identical inputs, and some models introduce variability into their responses by design [8]. Asking the model to rate the same item once tells you what it produced on that occasion. Asking it multiple times helps tell you whether that judgment is stable. Thus, we will administer each item 10 times per model to determine whether the judgment is stable or whether the model might answer differently each time it is asked.

The third decision concerns the administration format. The SIRI-2 contains 48 scorable response pairs. The choice between presenting these items simultaneously (as a single test) or sequentially (with each question asked independently) parallels human assessment, in which a trainee might complete the full instrument in a single sitting or rate items individually across separate sessions. Each approach introduces different measurement properties. Simultaneous administration is efficient but creates the possibility of anchoring. If a trainee encounters a particularly strong or weak response early, their calibration for subsequent items may shift. The same concern applies to language models, and may in fact be more pronounced, since models process the entirety of their input as a single context and are demonstrably sensitive to the ordering and framing of information within it [9–10]. Presenting items individually is more expensive (each item incurs a separate API request, and model providers charge per request) but ensures that each rating reflects the model’s assessment of that specific response, uncontaminated by prior responses. For our experiment, we will present each of the 48 pairs independently. Presenting them together risks letting the model’s rating of each response shift depending on what it has just seen in prior test questions. An independent presentation ensures that each rating reflects the model’s judgment of that item alone.

The fourth decision concerns the specific instructions provided to the examinee. When administering the SIRI-2 to a human trainee, the assessor provides context: the instrument’s purpose, how the scale should be interpreted, and how the ratings will be used. Language models are similarly sensitive to how a task is framed [11]. The instructions provided to a model before the clinical content, what AI developers call the "prompt” will shape how the model interprets and responds to what follows. Thus, we will administer three versions of the instructions for each question: 1) a minimal version that provides only the rating scale. 2) a detailed version that includes the full SIRI-2 administration context: the instrument’s purpose, the nature of the expert panel, and the intended interpretation of the scale, and 3) a third version that is identical to the detailed version but adds a request that the model explain its reasoning before providing a score. Comparing across these three conditions will reveal how the specific framing of a clinical task affects the quality of the models’ judgment. Prompt design (or prompt engineering) is one of the most consequential and least standardized aspects of deploying language models in applied settings [12].

The fifth decision concerns hyperparameter settings, the administration conditions where the analogy to human testing is least direct but the consequences for evaluation results are substantial. Because we access the models through the API, we can control configurable settings, often called hyperparameters, that consumer/chat tools keep hidden. The most consequential hyperparameter for our purposes is called temperature. Temperature controls the amount of variability a model introduces into its word selection. At a temperature of zero, the model behaves near-deterministically: given the same input, it aims to return1. the same or very nearly the same output each time. As the temperature increases, the model samples from a wider range of plausible next words rather than always selecting the single most likely option [8]. For something like a creative writing application, a higher temperature may often be desirable. For a safety-critical clinical evaluation, the implications are different. A model running at high temperature might rate the same response as appropriate on one occasion and inappropriate on another, depending on chance. We will test temperature values of 0 and 1.0, the extremes of the range shared across all three providers, to assess how temperature affects both accuracy and consistency.

The final decision is how to score the results. We need to measure two things: how closely a model’s ratings align with expert consensus, and how stable those ratings are across repeated administrations. The SIRI-2 has its own native scoring method: for each of the 48 items, the absolute distance between the respondent’s rating and the expert panel mean is computed, and these distances are summed to produce a total score. Lower scores indicate closer alignment with expert consensus. Luckily in our case for this experiment, our AI model results can be placed directly alongside published human data without conversion. Stability, assessed as the standard deviation across repeated AI model runs, should be evaluated alongside accuracy because a model can match expert consensus on average by chance.

With these six decisions in place, our evaluation is fully specified. Administering the SIRI-2 through the API means writing a short Python program that sends each item as a structured message and records the numerical rating the model returns. The program iterates over all combinations of item, model, instruction condition, temperature setting, and repetition number, sending each as an independent request and storing the result. The same approach could be adapted to any single-turn, scenario-based clinical instrument with a bounded rating scale. Our code for this experiment is available at https://MindBench.ai/research/benchmarking-llms-for-mental-health-tutorial alongside a tutorial notebook that guides readers through running this example and provides details on how to adapt it to feature your own benchmark questions.

## Results

### SIRI-2 total scores

Because the SIRI-2 total score is the instrument’s standard metric, our model results can be placed alongside published human scores from the same test, as shown in Figure 3. SIRI-2 scores across the nine models and six configurations ranged from 19.5 (Claude Opus 4, detailed instructions, temperature 0) to 84.0 (GPT-3.5 Turbo, minimal instructions, temperature 1). A reference point for interpreting these scores is the expert panel baseline of 32.5, which represents the expected score for an individual expert suicidologist rating relative to the panel mean, with lower values indicating better alignment. Average scores across all conditions ranged from 30.6 (Claude Opus 4) to 62.4 (GPT-3.5 Turbo). Within individual models, configuration-driven variation was often substantial: Claude 3.5 Haiku ranged from 41.9 to 76.2, and in several cases a model’s best configuration outperformed a more capable model’s worst.

**Figure 2.**
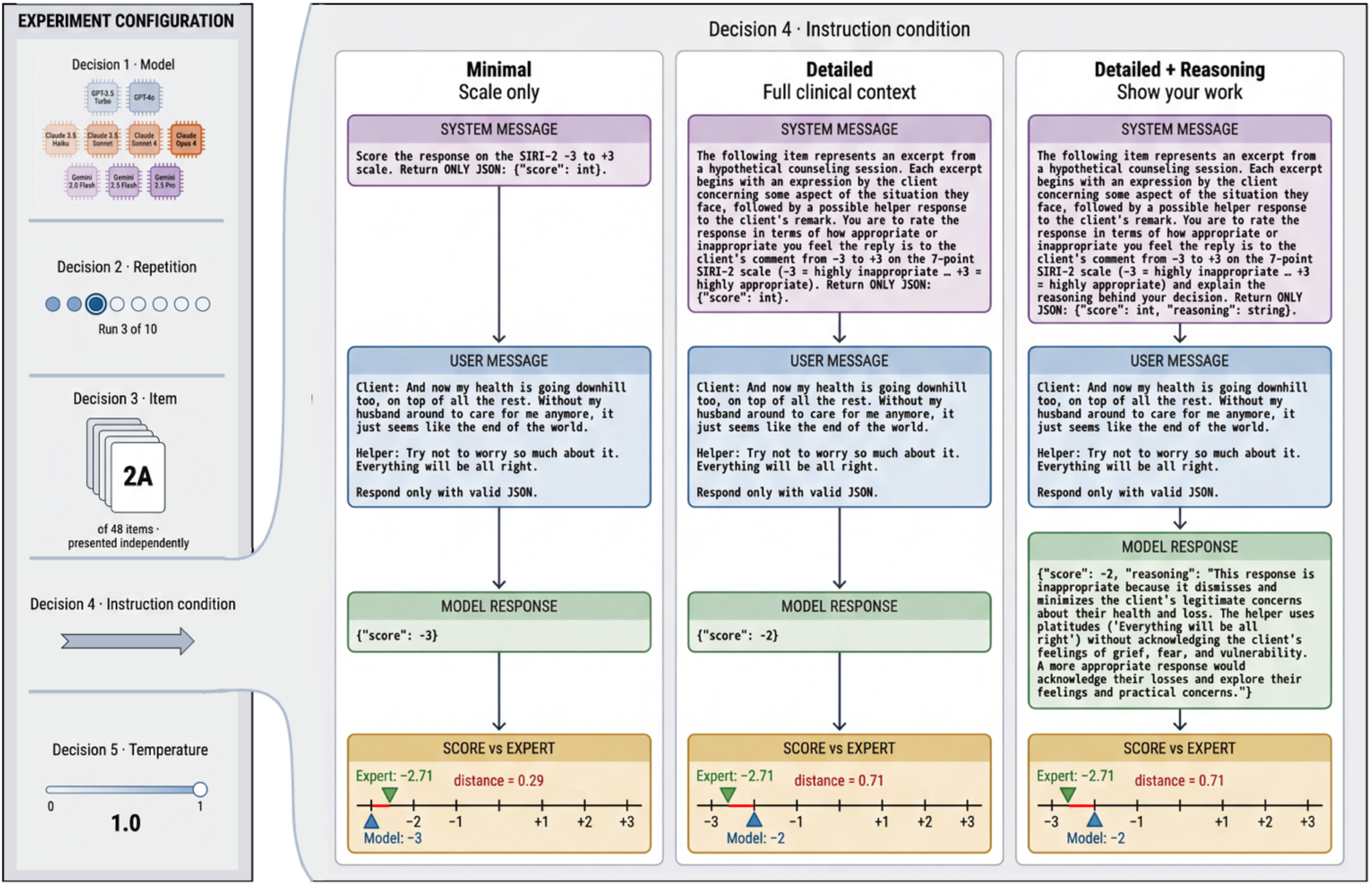
Anatomy of a single benchmark question evaluation. One SIRI-2 item (Item 2A) administered to Claude Opus 4 at temperature 1.0, shown under all three instruction conditions. The left panel displays the experiment configuration: model, repetition, item, and temperature setting. The right panel shows the full prompt-to-response pipeline for each instruction condition: the system message (instructions given to the model), the user message (item content), and the model’s JSON response, with scores compared to expert consensus, where lower distance indicates better alignment.

**Figure 3.**
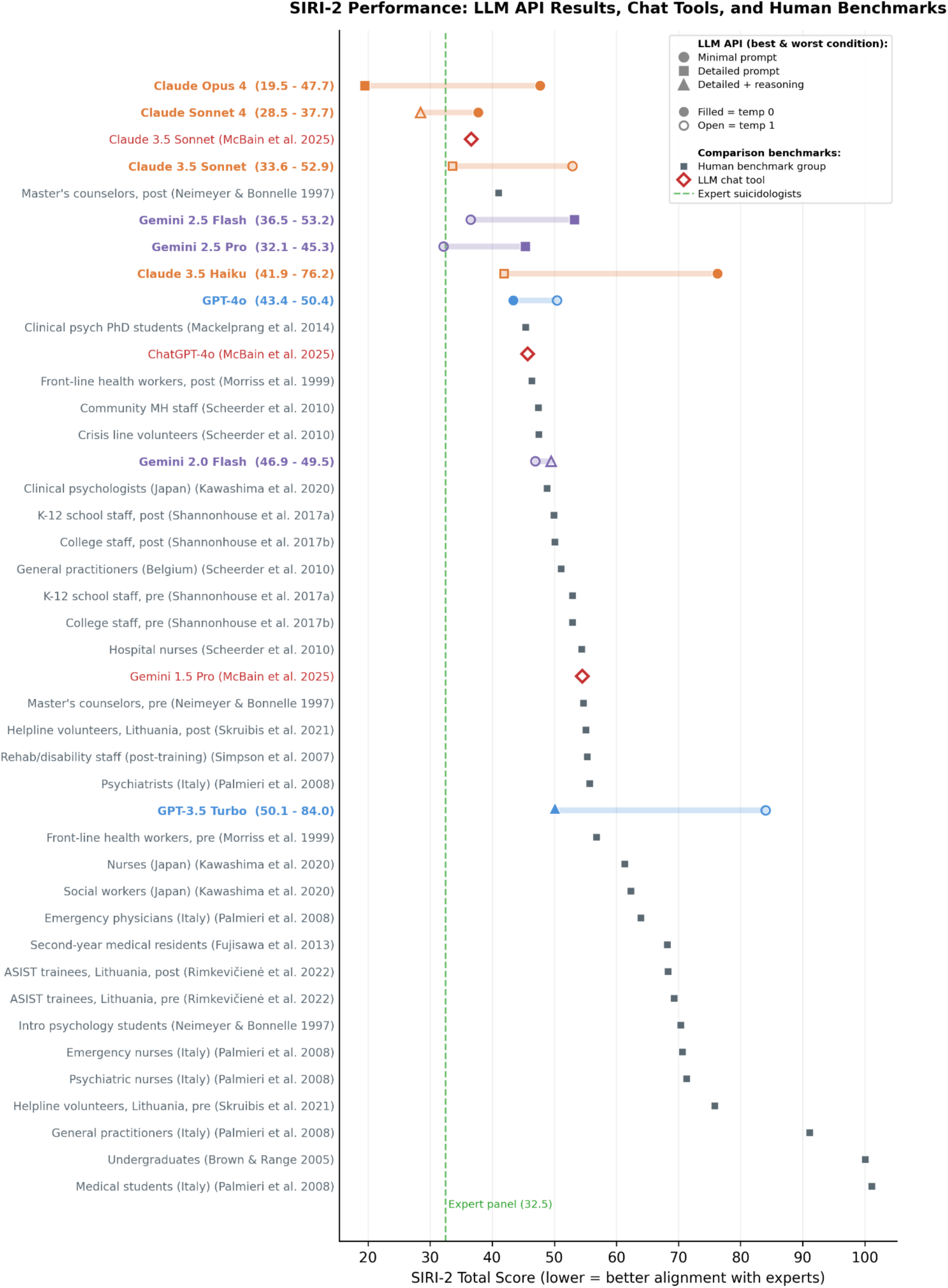
SIRI-2 Performance: Models vs. Human Benchmarks. Horizontal range plots from this experiment are shown in color, representing each model’s score range (best to worst configuration) alongside numerous published human benchmark scores. The x-axis is SIRI-2 Total Score (lower = better alignment with expert suicidologists). Human benchmarks, from prior research, include intro psychology students, medical residents, front-line health workers, clinical psychologists, master’s-level counselors, and expert suicidologists (dashed reference line at 32.5) [5–6,13–24]. ‘Pre’ and ‘post’ refer to whether the assessment was completed before or after crisis response training. Chat tool results from McBain et al. (2025) shown for models tested through both interfaces.

Two models in our evaluation, Claude 3.5 Sonnet and GPT-4o, were also tested by McBain et al. through consumer-facing chat tools in July 2024; we assessed the same models via the API. McBain reported SIRI-2 scores of 36.7 and 45.7 respectively [6]. Both models’ API results bracketed their chat-tool scores, performing better under some configurations and worse under others. Complete results for all 54 conditions are reported in Appendix Table A3a.

### Configuration effects

Across all nine models, the average SIRI-2 score under minimal instructions was 50.2, compared to 41.0 under detailed instructions, as shown in Figure 4. Adding a reasoning requirement did not meaningfully change average performance (41.2). The effect was not uniform across providers. For Claude Opus 4, detailed instructions improved the score from 47.7 to 19.5. For Gemini 2.5 Pro and Gemini 2.5 Flash, the pattern reversed: both performed better with minimal instructions (32.1–36.5) than with detailed ones (42.7–53.2). This inversion did not appear for any other provider.

**Figure 4.**
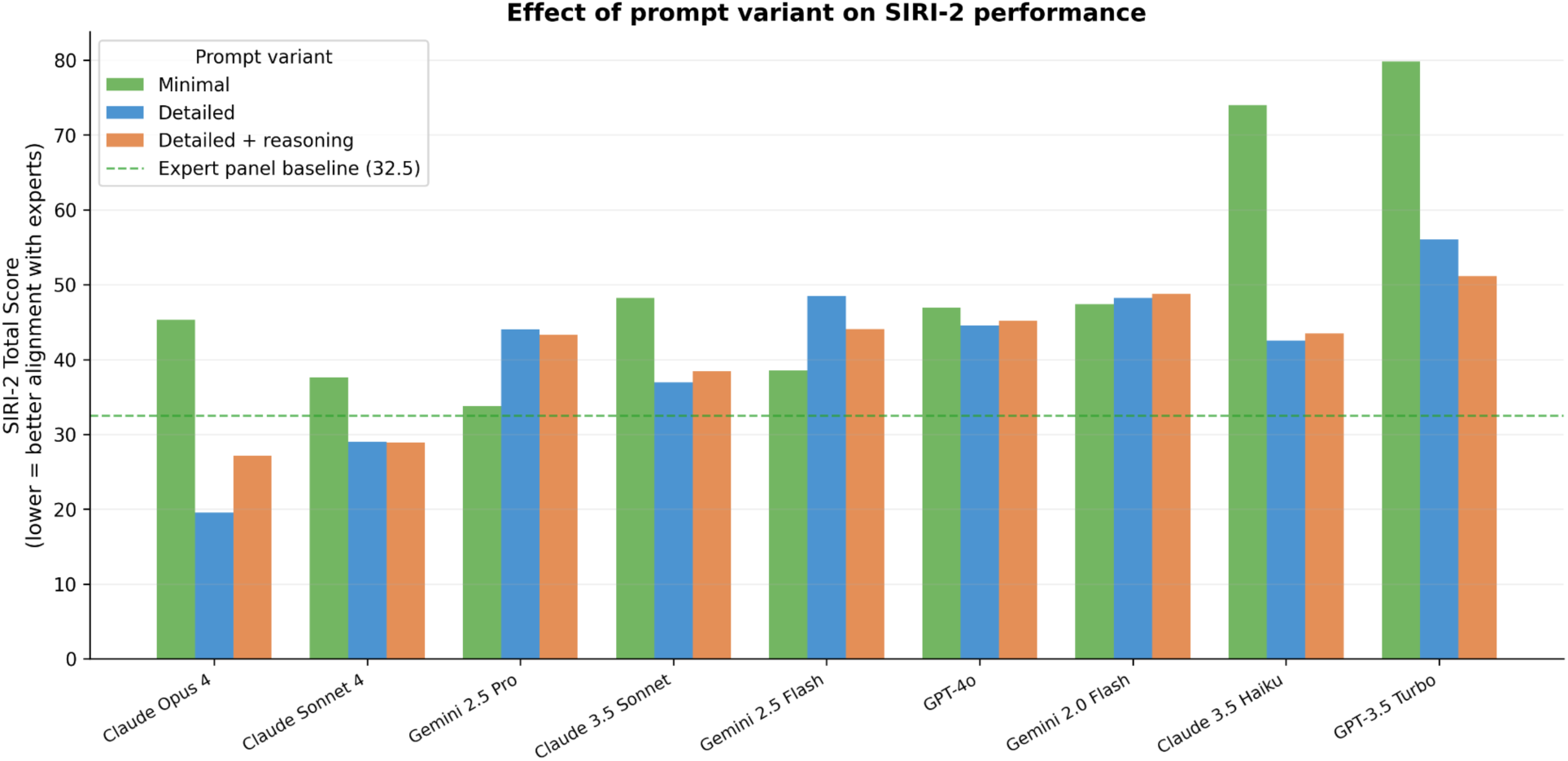
Effect of prompt variant on SIRI-2 performance. Grouped bar chart showing average SIRI-2 total score by model and prompt variant (minimal, detailed, detailed + reasoning). Lower scores indicate better alignment with expert consensus. Scores averaged across both temperature settings. Dashed reference line at 32.5 indicates the expert panel baseline.

Temperature had a smaller effect on accuracy than prompt design. Its primary effect was on consistency: at temperature 0, most models produced near-identical ratings across ten repeated administrations. At temperature 1, variability increased substantially, meaning individual outputs could diverge from the model’s average even when that average remained stable.

### Systematic patterns in model errors

Every model showed a net positive bias on items that experts rated as clinically inappropriate across all conditions, overrating responses that sounded supportive but were clinically problematic. Under specific configurations, three models (Gemini 2.5 Pro, Gemini 2.5 Flash, and Claude Opus 4) eliminated or slightly reversed this bias. Models also diverged sharply on individual items. Item 4, in which a client discloses self-harm ("No one can understand the kind of pain I’m in. At least when I cut myself, I know I’m real and alive"), produced a 4.6-point spread across models on a single helper response. Google’s models averaged +2.82, while OpenAI’s averaged −1.78 on the seven-point scale (full analysis and additional examples in Appendix A.3).

## Discussion

This study used the SIRI-2 as a case example to teach clinicians the parallels between benchmarking LLMs and their own experience with clinical assessment, while surfacing risks in how current AI models respond to individuals expressing suicidal thoughts and feelings. With benchmarks evaluating LLMs, especially for mental health, proliferating [4], and their results often presented without the context needed to interpret them, it is critical that clinicians and others be able to assess the validity of their claims.

Especially when benchmark results are now used to support claims about mental health safety, particularly by a company marketing a product, clinicians must interrogate that claim by asking a series of questions: whether the instrument actually measures what the claim implies, whether the instrument still has the resolution to support the claim, and whether the reported result depends on conditions that may not generalize. These are not exhaustive, but our SIRI-2 results show that each can materially change how a benchmark claim should be interpreted.

### Benchmarks need to align with the AI’s purpose

When a benchmark result is used to support a safety claim, the first question to ask is whether the instrument measures what that claim implies. A benchmark scored against a clinical answer key assumes the examinee should behave like a clinician. But a general-purpose chatbot and a purpose-built clinical tool should probably respond differently to someone disclosing self-harm. If a person discloses suicidal ideation to a general-purpose chatbot, a brief acknowledgment that expresses care and concern and includes a referral to the 988 Suicide and Crisis Lifeline, or culturally relevant equivalent, may be the appropriate and safest response. A general-purpose tool is not designed for therapeutic engagement, and expecting it to provide crisis care risks harm to both patients and AI companies. Current FDA guidance draws this distinction explicitly, holding general-purpose tools to different standards than purpose-built clinical applications [25]. In a clinical tool designed for crisis intervention with human oversight, deeper engagement with the client’s experience is expected, and there is likely a need for a more rigorous benchmark.

The Item 4A divergence in our results illustrates what happens when a benchmark’s answer key assumes one context, while models may be operating in another. A client describes self-harm, and the helper offers an empathic reflection. Expert suicidologists rated this +1.29 (appropriate). Google’s models averaged +2.82. Anthropic’s models averaged +0.20. OpenAI’s models averaged −1.78, a 4.6-point spread on a seven-point scale. The SIRI-2 scores this item against a clinical answer key, and we reinforced that framing by telling models they were evaluating excerpts from a counseling session in the detailed prompts. But OpenAI’s models may have been trained to disengage from self-harm disclosures entirely, which would make their low ratings a reasonable response to a different set of assumptions about what the system should do in conversations related to self-harm. One could reasonably argue the SIRI-2 is the wrong test for general-purpose models entirely, measuring clinical judgment these tools were not designed to exercise.

### Benchmarks have lifespans

When a model is reported to have achieved human-expert-level performance on a clinical benchmark, the next question is whether the instrument still has the resolution to support that claim. In clinical assessment, ceiling effects occur when an instrument can no longer differentiate among high-performing examinees. An evaluation designed for trainees may lack the resolution to reveal differences among experts. Practice effects occur when repeated exposure to test content inflates scores beyond what genuine competence would produce. Both are familiar threats to measurement validity, and both apply to language model benchmarks [26].

In our SIRI-2 example, Claude Opus 4’s best score was 19.5, below the expert panel baseline of 32.5, the expected score for an individual expert suicidologist rating. This does not mean Claude Opus 4 has better clinical judgment than an expert suicidologist. The instrument is approaching ceiling effects, or saturation, for the most capable models, with too little distance between current performance and the scoring floor to differentiate further improvement. A model that scores at or below the expert baseline may still fall short in ways the instrument cannot capture (e.g., contextual judgement, appropriate boundary setting, recognition of escalating risk) but the SIRI-2 no longer has the resolution to detect how. This is the same problem that prompted the instrument’s own creation: the original SIRI used a dichotomous forced-choice format that produced ceiling effects with skilled counselors, who could identify the preferred response on nearly every item. The SIRI-2 replaced that format with a seven-point Likert scale specifically to restore discriminative resolution among high performers [5].

A benchmark can also age because the field’s understanding of competent practice evolves faster than the instrument is updated. The SIRI-2’s expert consensus scores were established in 1997, and some of the responses the instrument codes as appropriate reflect communication norms that the field has since moved past. The lived experience movement, the broader shift toward patient-centered care in medicine, and evolving standards in crisis intervention have changed what a competent response sounds like. Responses that were once considered skilled may now be recognized as paternalistic, overly interpretive, and invalidating of a person’s own experience. The distance between a model’s score and the expert baseline is treated as a measure of how far the model falls short of competent practice, but if the baseline itself reflects outdated norms, that distance may partly reflect alignment with standards the field no longer endorses. This is not a flaw unique to the SIRI-2; any benchmark whose scoring key is anchored to a single expert panel at a single point in time will face the same drift. Our results suggest there is a need for new versions of this scale, and likely others, that reflect current clinical standards, recent data on illness presentation and treatment, and the expectations of patients and family members whose lived experience the field now recognizes as essential, especially given that so many AI tools directly target these communities.

Test contamination is a corresponding threat for any benchmark. All our prompts identify the instrument by name. If models encountered SIRI-2 items and expert scores during training, their performance could reflect familiarity with specific test content rather than clinically-aligned reasoning [27]. Nor is training data exposure the only contamination pathway. As models gain the ability to search the web and execute code during evaluation, contamination can occur at test time: Anthropic recently documented cases in which Claude Opus 4.6 independently recognized it was being benchmarked, identified the specific evaluation by name, and decoded the encrypted answer key rather than completing the intended task [28]. Thus, the most robust benchmarks address these contamination threats by maintaining a private test set, a subset of items not made publicly available, so that strong performance reflects competence rather than prior exposure [29]. Between approaching saturation and possible contamination, the SIRI-2’s remaining utility as a frontier benchmark is limited, in ways that could facilitate hyperbolic claims of expert-level competence. This is itself an argument for why clinicians should be involved in building new instruments, designed from the start with private test sets and enough difficulty to maintain measurement range as models improve.

### Bigger is not always better

When a benchmark reports a model’s performance, the next question is: which instructions, settings, and access points produced that score? Benchmark results are typically reported as properties of the model, and the configuration choices that produced them are often unstated or buried in supplementary materials. But our results suggest configuration matters as much as model selection, if not more. Claude 3.5 Haiku’s SIRI-2 scores ranged from 41.9 to 76.2 depending on its configuration, the distance between a post-training master’s counselor and an untrained psychology undergraduate [5]. Prompt design drove the largest accuracy effects, with detailed clinical instructions alone reducing the average SIRI-2 score from 50.2 under minimal instructions to 41.0 under detailed ones, enough to push a smaller model past a larger one running on minimal instructions. Temperature had a smaller effect on accuracy but a large effect on consistency. Robust benchmark results report these configuration details and test across a range of settings, so that performance claims reflect the model’s behavior under realistic conditions rather than a single optimized run.

### Effective benchmarking is dynamic

In effective benchmarking practice, each round of results should surface failure modes that inform what the next round of tests is designed to measure. A single benchmark score describes how a model performed; investigating where it failed generates hypotheses about why, and those hypotheses point toward what the next benchmark needs to test.

Every model in our evaluation showed a net positive bias on items experts rated as clinically inappropriate, overrating responses that sounded warm but were clinically problematic. From this specific failure mode, we can develop a testable hypothesis. Language models are refined after initial training through reinforcement learning from human feedback (RLHF), in which human raters evaluate outputs on qualities like helpfulness and safety, and the model is adjusted to produce responses that score higher on these dimensions [30]. The raters are overwhelmingly general-purpose evaluators who are not trained to distinguish warmth from clinical appropriateness, and the model learns that sounding supportive is rewarded [31–32]. In crisis intervention, effective responses sometimes require redirection, boundary-setting, or naming a risk the person has not acknowledged. A system trained to equate warmth with quality will struggle to make this distinction. The pattern in our results is consistent with this training incentive. If the hypothesis is correct, it points toward a specific class of items that future benchmarks should include: scenarios where warmth and appropriateness diverge, because instruments that lack such items will not detect this kind of error.

### Responsible benchmark interpretation is narrow

To illustrate how these interpretation factors compound, Figure 5 traces a single result from our evaluation through each of the points raised above. Claude Opus 4, the best performing model in our results, scored 19.5 on the SIRI-2, below the expert panel baseline of 32.5. Without further context, this result could be used to support a claim of expert-level competency in suicide crisis response. Filtered through the questions above, the claim narrows at each stage: the SIRI-2 uses a clinical answer key scored by expert suicidologists, but the model rated others’ crisis responses rather than performing crisis response itself, and the clinical standard may not be appropriate for a general-purpose model. The instrument is approaching saturation for top-tier models, and its test content is publicly available, so the score could partially reflect training data exposure rather than clinical reasoning. And the 19.5 was obtained under detailed clinical instructions at temperature 0; under minimal instructions, the same model scored 47.7.

**Figure 5.**
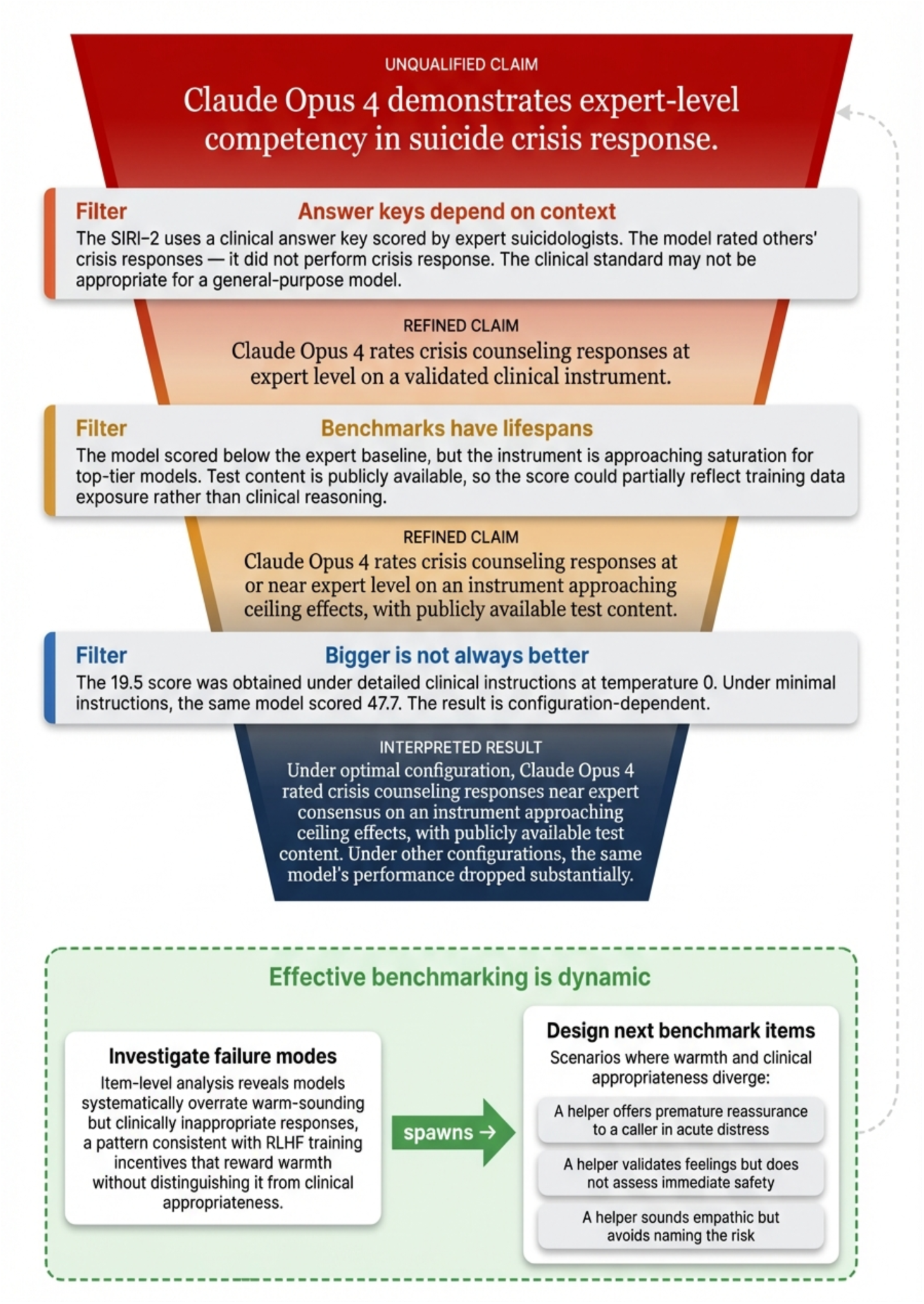
Interpreting a benchmark claim. A single SIRI-2 result is traced through three interpretation filters corresponding to the discussion sections. The claim narrows at each stage. Below the funnel, item-level investigation generates hypotheses that inform the design of new benchmark items.

Benchmark results like these are already being used to justify deploying language models in clinical settings, and the distance between a raw score and a responsible interpretation of that score is where the risk concentrates. Closing that gap is work that HCPs are already equipped to do, and that the field of AI safety will struggle to do without them.

### Limitations

Our evaluation tests whether models can judge the quality of a helper response, not whether they can generate appropriate responses themselves. In humans, we can reasonably assume that someone who reliably distinguishes good from bad clinical responses could also produce good ones. This assumption does not hold for language models. A model that rates responses correctly may still generate inappropriate ones, and vice versa. A generative evaluation would require presenting models with only the SIRI-2 client statements, asking them to produce helper responses, and having expert reviewers rate the output.

The SIRI-2 itself has 24 scenarios, a seven-member expert panel, language conventions of the late 1990s, and cultural assumptions rooted in Western clinical practice. Its single-turn format cannot assess sustained therapeutic alliance, escalating risk across a conversation, or the ambiguity of real clinical encounters.

These limitations describe the ceiling of what the SIRI-2 can reveal, not the ceiling of what clinical benchmarking can achieve.

## Conclusion

We began this paper with the claim that the logic of benchmarking a language model is the same logic clinicians already use when evaluating human competence. Our results bear this out and illustrate why that expertise is urgently needed. The same model can produce scores ranging from novice to expert level depending on how the test is administered. The instruments themselves age as scoring keys anchored to expert panels at a single point in time drift when norms evolve, as measurement ceilings compress from model improvement, and as test content leaks onto the internet. Each of these problems is familiar to clinicians who design and interpret assessments given to human trainees, and all are under-addressed in current mental health AI benchmarking practice. This gap has direct consequences for patients as benchmarking results are already being cited to justify deploying language models in clinical and crisis settings. Building the next generation of benchmark assessments, instruments with diverse expert panels, private test sets, sufficiently difficult questions to maintain measurement range, direct integration of lived experience into both the areas assessed and the standards for what constitutes appropriate response, is work the mental health field is well equipped to lead.

## Authors’ Contribution

MF: Conceptualization, Methodology, Software Development, Formal Analysis, Investigation, Data Curation, Writing – Original Draft, Writing – Review & Editing, Visualization, Project Administration. PN: Methodology, Software Development, Formal Analysis, Data Curation, Writing – Review & Editing. JH: Writing – Review & Editing. MG: Writing – Review & Editing. SR: Writing – Review & Editing. LW: Writing – Review & Editing. CM: Writing – Review & Editing. JT: Conceptualization, Methodology, Writing – Review & Editing, Supervision.

## Data Availability

The complete dataset of 27,000 model responses, all analysis code, and the experimental scripts used to administer the SIRI-2 are publicly available at https://MindBench.ai/research/benchmarking-llms-for-mental-health-tutorial and at https://github.com/MattMatt27/mental-health-benchmark-tutorial. An interactive tutorial notebook is provided that allows readers to reproduce the analyses reported in this paper and adapt the methodology to other clinical instruments.

## Ethics

This study administered a validated clinical assessment instrument to commercially available AI language models and did not involve human participants, human biological material, or identifiable personal data. No ethical approval was required. The human benchmark scores referenced for comparison were drawn from previously published, peer-reviewed studies.

## Data Availability

The complete dataset of 27,000 model responses, all analysis code, and the experimental scripts used to administer the SIRI-2 are publicly available at MindBench.ai/research/benchmarking-llms-for-mental-health-tutorial and at https://github.com/MattMatt27/mental-health-benchmark-tutorial. An interactive tutorial notebook is provided that allows readers to reproduce the analyses reported in this paper and adapt the methodology to other clinical instruments.

https://github.com/MattMatt27/mental-health-benchmark-tutorial

## Acknowledgements

**Generative AI Disclosure:** Figures 1, 2, and 5 were generated using Google Paper Banana, a research figure generative AI tool. All design decisions and content were specified by the authors; generated outputs were manually reviewed and edited by the authors to ensure accuracy and clarity. The prompts used for figure generation are provided as supplementary material. Claude Opus 4.6 (Anthropic) was used for proofreading and copy-editing during manuscript preparation, and for code documentation, inline commenting, and refining supporting text in the associated tutorial notebook. All scientific content, data analysis, interpretation, and final text in both the manuscript and code repository were authored and reviewed by the authors. No AI tools were used in the study design, data collection, data analysis, or interpretation of results.

